# Delays during PBMC isolation have a moderate effect on yield, but severly compromise cell viability

**DOI:** 10.1101/2022.01.02.22268625

**Authors:** T Golke, P Mucher, P Schmidt, A Radakovics, M Repl, P Hofer, T Perkmann, M Fondi, KG Schmetterer, H Haslacher

## Abstract

**Background:** Peripheral blood mononuclear cells (PMBCs) are a versatile material for clinical routine as well as for research projects. However, their isolation via density gradient centrifugation is still time-consuming. When samples are taken beyond usual laboratory handling times, it may sometimes be necessary to pause the isolation process. Our aim was to evaluate the impact of delays up to 48 hours after the density gradient centrifugation on PBMC yield, purity and viability.

**Methods:** PBMCs were isolated from samples of 20 donors, either with BD Vacutainer CPT tubes (CPT) or with the standard Ficoll method. Isolation was paused after initial density gradient centrifugation for 0, 24, or 48 hours. PBMC yield, purity and viability were compared.

**Results:** The yield did not change significantly over time when CPT were used (55%/52%/47%), but did after isolation with the standard method (62%/40%[p<0.0001]/53%[p<0.01]). Purity was only affected if CPT were used (95%/93%[p=n.s./92%[p<0.05] vs. 97% for all time points with standard method). Whereas viable PBMCs decreased steadily for CPT isolates (62%/51%[p<0.001]/36%[p<0.0001]), after standard Ficoll gradient isolation, cell apoptosis was more pronounced already after 24h delay, and viability did not further decrease after 48h (64%/44%[p<0.0001]/40%[p<0.0001]).

**Conclusions:** In conclusion, our data suggests that post-centrifugation delays of up to 48h might have only a minor effect on cell yield and purity. However, at the same time, a relevant decrease in cell viability was observed, which could be partially compensated by the use of CPT if the isolation was resumed latest the day after blood withdrawal.

## Introduction

Peripheral blood mononuclear cells (PBMCs) are employed to investigate a broad range of diagnostic and research-related questions (1-4). The cells, which represent a diverse fraction of white blood cells including lymphocytes and monocytes, are usually separated in a density gradient (5). This step is time consuming, as it comprises the density gradient centrifugation, which is usually performed with low a- and deceleration, as well as several washing and preservation steps.

The original method included the layering of diluted, anticoagulated whole blood on density fluids, as e.g. Ficoll or Percoll (6, 7). Standardization and simplification efforts lead to the development of specific tubes, which – besides the density fluid – contain physical barriers that separate the PBMCs from other cellular blood components after centrifugation. In BD Vacutainer® Cell preparation tubes (CPT; Becton Dickinson, Franklin Lakes, USA), blood can be drawn directly, hence, no additional layering is necessary (8).

As mentioned above, the isolation of PBMCs is despite all standardization still time consuming and should be performed soon after blood withdrawal (9). Biobanks, which are facilities that specialize in the handling, storage, and distribution of biomaterials and related data, often receive samples for PBMC isolation throughout the day if some of their collections include viable cells. However, PBMC isolations of samples that are received later in the day can sometimes not be completed on the same day without working overtime. Manufacturers of isolation tubes recommend in this case to perform the density centrifugation step only and to continue with the washing steps at a later time point. Unfortunately, no data is so far available on the impact of this approach on the yield, the purity and the viability of PBMCs.

We, therefore, aimed to investigate the impact of delays up to 48 hours after the density gradient centrifugation step on PBMC quality. PBMCs were either isolated with CPT or, as a control, by standard Ficoll layering. Moreover, we evaluated the effect of different storage temperatures during these delays.

## Methods

### Study design and participants

Blood of 20 participants of the MedUni Wien Biobank “Healthy Donor Cohort” was included into this prospective method evaluation study. Inclusion criteria were an age ≥18 years and willingness to give written informed consent, whereas insufficient biomaterial would have led to exclusion from the study. All donors provided written informed consent (ethics committee No. 404/2012). The study protocol is compliant with the Declaration of Helsinki and was reviewed and approved by the ethics committee of the Medical University of Vienna (No. 1022/2021). All samples were processed and stored by the MedUni Wien Biobank, a central facility with certified quality management (ISO 9001:2015) (10).

In brief, each participant donated 49 mL of blood. 1mL was drawn in EDTA-tubes and used to perform a total blood count, 24mL were collected in 3 BD Vacutainer CPT tubes (Becton Dickinson, Franklin Lakes, USA), and the remaining 24mL in 3 lithium heparine tubes (Greiner Bio-One, Kremsmuenster, Austria) for later PBMC isolation. The first isolation step (densitiy gradient centrifugation) was performed within one hour after blood collection. Then, one tube was used for each patient and isolation method to wash, aliquot and store cells either immediately (0h), after a 24h or an 48h delay, were cells were kept on 2-10°C. For five 5 donors, drawn CPT tubes were kept on 2-10°C and on room temperature after the density gradient centrifugation step to exclude a possible influence of post-centrifugation storage temperatures. A flow chart is presented in figure 1.

**Fig. 1:**
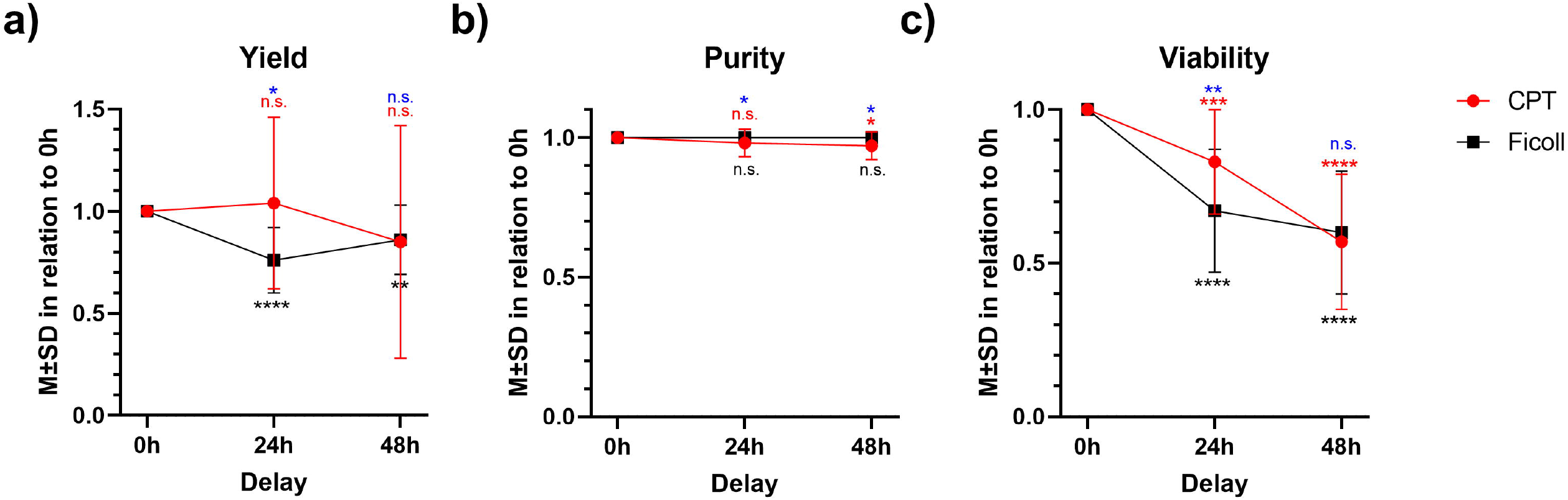
PBMC yield (a), purity (b), and viability (c) measured after a 24h or 48h delay in PBMC isolation protocols were compared to undelayed isolates (0h). Red/black asterisks indicate significant deviations at the respective delay (24h, 48h) from the undelayed sample (0h) for CPT/standard Ficoll gradient centrifugation protocols. Blue asterisks mark the differences between isolation methods at a given delay. M±SD… mean±standard deviation; *… p<0.05, **… p<0.01, ***… p<0.001, ****… p<0.0001, n.s…. not significant.

### Blood withdrawal and PBMC isolation

Blood was drawn either directly into cell preparation tubes (CPT) or in lithium heparine tubes for Ficoll density gradient centrifugation. An additional small amount was used for a total blood count on an XN-2000 Hematology Analyzer (Sysmex, Kobe, Japan).

CPT tubes were centrifuged at 1,800xg for 20 minutes at room temperature. For isolation according to a standard Ficoll density gradient centrifugation protocol, blood was diluted 1:2 in phosphate buffered saline (PBS; Carl Roth, Karlsruhe, Germany) and centrifuged at 863xg for 20 minutes at room temperature, without a fast start and a break. The cell layer was transferred to a sterile 15mL tube and 10mL of PBS was added. The washing steps described below were performed either immediately, after 24h or after 48h. For the latter two time points, CPT were meanwhile stored at 2-8°C as recommended by the manufacturer.

If this has not already been done yet (CPT), cells were transferred to a sterile 15mL tube and washed twice with at 311xg, 15 min, room temperature. After the washing steps, cells were reconstituted in a freezing medium consisting of 70% Iscove’s Modified Dulbecco’s Media inclusive Gentamycin (IMDM; Gibco, Grand Island, USA), 20% fetal bovine serum (Pan-Biotech, Aidenbach, Germany) and 10% dimethyl sulfoxide (DMSO; Carl Roth). A small portion of each isolate was used for the assessment of yield and purity. The remaining cell solution was transferred to cryovials which were slowly frozen for 48 hours in Mr. Frosty containers (Thermo Scientific, Waltham, USA) at <-70°C and then moved to LN_2_ until viability assessment.

### Assessment of yield and purity

As described above, total cell counts were performed on both the whole blood samples, as well as the final cell isolates of each donor. Yield was defined as the proportion of the isolated PBMCs (lymphocytes and monocytes) counted before freezing to the lymphocytes and monocytes that were present in the 8mL of whole blood used: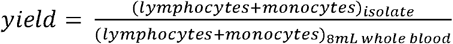. Purity was defined as the proportion of PBMCs to all leukocytes counted after isolation: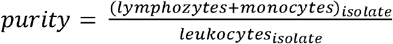. All analyses were performed on an XN-2000 Hematology Analyzer (Sysmex).

### Viability staining

Cryovials were taken from LN_2_ and thawed at room temperature. Then, samples were centrifuged at 300xg for 10 minutes at room temperature. After discarding the freezing media, cells were washed with 10mL HBSS (Gibco) and cell viability was assessed using a commercially available Annexin V Apoptosis Detection kit (Thermo Scientific). In brief, cells were washed once with 2mL of 1x Binding buffer at 330xg for 10 minutes, room temperature. The cell pellets were then reconstituted in 100µL Binding Buffer + 2µL Annexin V APC. After an incubation step (15 minutes, room temperature, light protected), cells were again washed with 1mL 1x Binding Buffer. Subsequently, 100µL Binding Buffer + 1µL propidium iodide were added to the cell pellets. Flow cytometry was performed on a BD FACSCanto II and results were analysed using the FACSDiva software (Becton Dickinson). As apoptotic cells start to incorporate Annexin V and, at a later stage, also propidium iodide, only cells that were both Annexin V and propidium iodide negative were considered viable.

### Statistical analysis

If not otherwise indicated, continuous data are given by means±standard deviation. Main effects and the interaction of post-centrifugation delays and isolation methods (CPT vs. Ficoll) were evaluated by two-way repeated-measures ANOVAs with two within-subject variables. Post-hoc comparisons were performed according to Sidak. Paired data with groups sizes ≤10 were compared by Wilcoxon tests. Results with p-values <0.05 were considered significant. All calculations were done with SPSS 23.0 (IBM, Armonk, USA) and graphs were drawn using GraphPad 7 (Graph Pad Inc., La Jolla, USA).

## Results

### Impact of isolation delays on PBMC yield

As described above, PBMCs from 20 healthy donors were either isolated using CPT or by a standard Ficoll density gradient centrifugation protocol, whereby isolation was paused for 0h, 24h or 48h, respectively, after the initial centrifugation step. After extraction, the PBMC yield was determined.

For isolates for which extraction was completed on the day of blood withdrawal (0h delay), mean yields were 55±17% for CPT and 62±9% after standard Ficoll isolation. 24h/48hs delay after the first centrifugation step reduced the yield to 52±12%/40±20% in CPT and to 47±11%/53±10% applying Ficoll layering. A two-way repeated-measures ANOVA with two within-subject factors (delay and tube type) revealed a significant main effect for the isolation delay (F=11.8, df_1_=2, df_2_=38, p<0.001) and a significant interaction delay x tube type (F=3,6, df_1_=2, df_2_=38, p=0.036), indicating a different effect of isolation delays on both methods. In Sidak-corrected pair-wise comparisons, only the differences between isolates derived from different delays during Ficoll gradient centrifugation remained statistically significant.

To visualize the deviations resulting from an isolation delay more clearly, the yields from the delayed isolations were subsequently presented as a percentage of the yields of the undelayed isolation (Fig. 1a). For CPT, PBMC yields did not significantly decline over time, although a considerable variability was recorded (ranges after 24h/48h delay: 26-213%/10-194% compared to 0h delay). After standard Ficoll gradient isolation, in contrast, PBMC yields were significantly lower after both 24h (76±15%) and 48h (86±17%) delay when compared to baseline.

Hence, it appears as if delays in isolation protocols for up to 48 hours have a less significant impact when CPT are used. In the next step, the effect on the purity of PBMC isolates was examined.

### Impact of isolation delays on PBMC purity

For the present study, PBMC purity was defined as the share of monocytes and lymphocytes in all isolated leukocytes. Mean purities were above 90% for all isolates (0h/24h/48h, CPT: 95±3%/93±6%/92±6%, Ficoll: 97±1%/97±2%/97±1%). In a two-way repeated-measures ANOVA, the isolation method (CPT or Ficoll) presented with a significant main effect (F=11.3, df_1_=1, df_2_=19, p=0.003), indicating a purer cell isolate after the Ficoll method (difference, mean±SEM: -4±1%). A significant interaction isolation method × delay (F= 5.2, df_1_=2, df_2_=38, p=0.10) also suggests a difference in the effect of delays for the individual isolation methods. However, post hoc Sidak-corrected tests did not produce any statistically significant results.

Differences from baseline purities (set 100%) were plotted for both isolation methods, and again, results from CPT tubes presented with greater variability (ranges after 24h/48h delay for CPT: 85-103%/84-104% and for Ficoll 97-104%/97-103%, both compared to 0h). However, only the purity after a 48h delay in CPT tube isolation significantly differed from the purity of the undelayed isolate (Fig. 1b).

These results suggest that PBMC purity, which was significantly higher after standard Ficoll gradient centrifugation than with CPT tubes, was only marginally affected by delayed isolations. Finally, cell viability was assessed.

### Impact of isolation delays on PBMC viability

PBMC viability was assessed after storage in LN_2_ using a commercially available Annexin V/propidium iodide staining kit. Only cells without propidium iodide and Annexin V signals in flow cytometry were classified as viable. In the two-way repeated-measures ANOVA, the frequency of both Annexin V and propidium iodide negative, i.e., viable, cells declined with increasing delay (F=70.9, df_1_=2, df_2_=38, p<0.0001) from (mean±SEM) 63±2% (0h) to 47±3% (24h) and 38±3% (48h). However, besides this significant main effect, the interaction isolation method x delay was significant as well, indicating a different impact of delays on the isolation methods (F=6.8, df_1_=2, df_2_=38, p=0.003). Whereas viable PBMCs decreased steadily for CPT isolates, after standard Ficoll gradient isolation, cell apoptosis was more pronounced already after 24h delay, and viability did not further decrease after 48h. In CPT tubes, only 83±17%/57±22% of the PBMCs that were viable in undelayed samples lacked apoptosis signals after 24h/48h. After standard Ficoll gradient centrifugation, cell viabilities compared to the undelayed samples were 67±20%/60±20% after 24h/48h (Fig. 1c).

Taken together, our results suggest that despite minor reductions in cell yield and purity, delays during PBMC isolations considerably affect PBMC viability.

### The influence of storage temperature after density gradient centrifugation

However, several protocols suggest to keep CPT tubes rather on room temperature instead of 2-10°C to prevent platelet activation and apoptosis. To quantify the influence of storage temperature after the first centrifugation, we repeated the experiment with sample material from ten (yield/purity) or five (viability) different samples using only CPT tubes.

In Fig. 2, yield represents the proportion of PBMCs that could be successfully isolated from the primary material (a), purity represents the proportion of PBMCs within the isolate (b), and viability represents the proportion without Annexin V/propidium iodide uptake (c). In Wilcoxon analysis, samples kept at room temperature presented with a higher yield (24h: +0.16, p=0.013; 48h: +0.14, p=0.037), but with a lower purity, especially after the 24h centrifugation delay (24h: - 0.01, p=0.017; 48h: -0.01, p=0.093). However, storage at 2-10°C was not inferior to room temperature at 24h (+0.01, p=0.893) and resulted in an even better viability after 48h (+0.28, p=0.043).

**Fig. 2:**
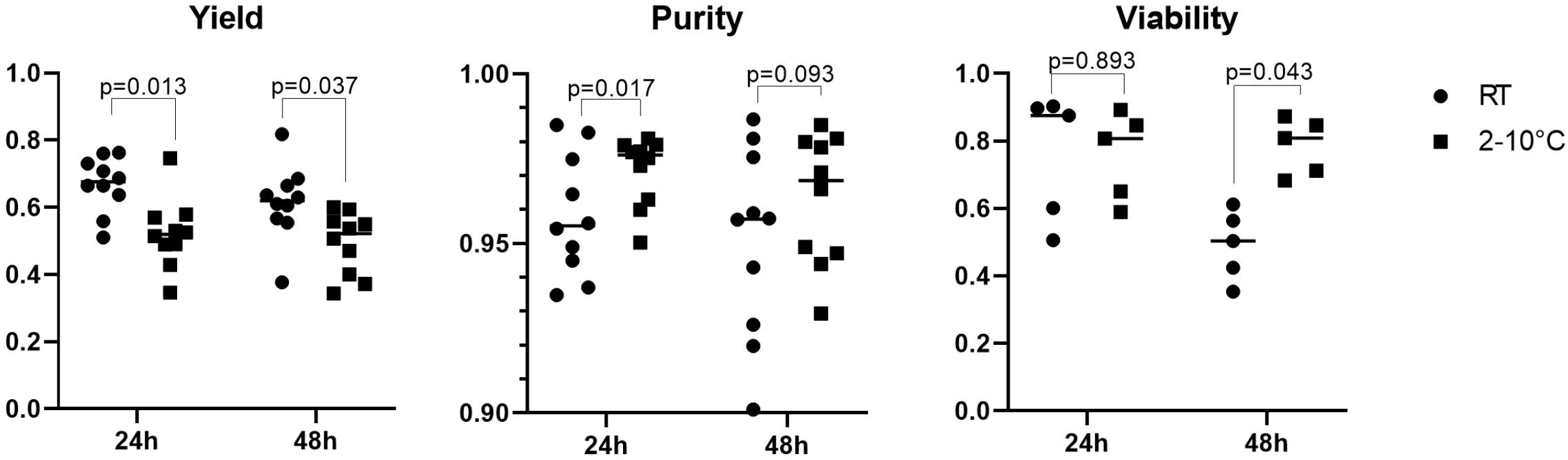
Differences in yield (n=10 samples per group), purity (n=10 samples per group) and viability (n=5 samples per group) after 24h and 48h post-centrifugation delays in samples stored at room temperature (RT, filled circles) vs. 2-10°C (filled squares).

## Discussion

Post-centrifugation delays during the time-consuming PBMC isolation are often inevitable in pre-analytical facilities that do not operate on a 24/7 basis. Although delays up to 48h might not relevantly reduce cell yield if special cell preparation tubes are used, cell viability is significantly affected. To the best of our knowledge, this phenomenon has not been systematically reported before.

There is certain evidence that the time to the first centrifugation step is critical regarding cell yield, purity and viability. PBMC counts after a pre-isolation delay of 30 hours when compared to cells extracted within the first day after blood withdrawal (11). Regarding purity, an 84-fold increase in granulocyte contamination was reported when whole blood was refridgerated for 22-26 hours before PBMCs were isolated (12). Cell viability was shown to be relevantly decreased if isolation started 24 hours after blood withdrawal in both CPT and Ficoll to tubes, although it has to be mentioned that only samples from three donors were included (13).

Reports regarding the effect of post-centrifugation delays can be hardly found. A recent review presented data of only three samples (9), which are, indeed, in line with our observations. Our experimental setup differed regarding the temperature, at which samples were stored after centrifugation. Although protocols suggest to keep samples at room temperature, we decided to refridgerate centrifuged CPT tubes at 2-10°C during the post-centrifugation delay to ensure comparability with standard-Ficoll preparations, which were refridgerated as well. This was done in order to curb the cell metabolism, as Ficoll poses a potentially toxic environment for cells (14) and alters their electrolyte and trace element composition (15). To assess, whether our findings were solely attributable to the post-centrifugation storage temperature, we compared samples stored at room temperature to those stored at 4°C after the first centrifugation step. Although the latter showed a slightly lower yield, they were significantly more pure and had a lower platelet contamination. Moreover, storage at 2-10°C instead of room temperature did not seem to further impair viability – in fact, the proportion of viable cells after an 48h centrifugation delay was significantly higher in refrigerated samples (Fig. 2).

In conclusion, our data suggests that post-centrifugation delays of up to 48h, as permitted by manufacturers of PBMC preparation tubes, might have only a minor effect on cell yield and purity. However, at the same time, a relevant decrease in cell viability was observed, which could be partially compensated by the use of CPT if the isolation was resumed latest the day after blood withdrawal. Thus, it appears that both biobankers and researchers must be aware that the logistic advantage of delaying PBMC isolations after the density gradient centrifugation might come at the cost of cell viability.

## Data Availability

All data produced in the present study are available upon reasonable request to the authors.

## Acknowledgments

We thank all participants for their valuable donations. The MedUni Wien Biobank is part of the Austrian biobanking consortium BBMRI.at.

## Research funding

None.

